# Open Dialogue compared to usual care for adults experiencing a mental health crisis: A feasibility study for the ODDESSI trial

**DOI:** 10.1101/2025.10.28.25332665

**Authors:** Stephen Pilling, Kirsty James, Katherine Clarke, Thomas Craig, Shaira Hassan, Yasmin Ishaq, Catherine Kinane, James Osborne, Georgie Parker, Russell Razzaque, Sabine Landau

**Author notes:** Corresponding author: Stephen Pilling. Joint first author (Stephen Pilling, Kirsty James). Trial registration: ISRCTN17977792. Ethical Approval: REC Reference: 18/LO/0868, IRAS Project ID: 233243. Funding: Funding for this research programme was provided by the National Institute for Health Research (NIHR) Programme Grant for Applied Research Project Number: RP-PG-0615-2002.

## Abstract

**Background:** Open Dialogue is a complex mental health intervention which places strong emphasis on working with service users’ social network in conjunction with provision of crisis and community care. It is both an organisational intervention of the mental health team and a clinical intervention at the individual level. This study examines the feasibility of conducting a cluster randomised controlled trial to evaluate the clinical and cost-effectiveness of Open Dialogue compared to Usual Care in the English National Health Service.

**Methods:** Coterminous GP practice clusters were randomised to Open Dialogue or Usual Care in preparation for a future evaluation trial. The cluster randomisation approach used restricted randomisation to balance intervention groups for catchment area of the community mental health team, GP practice size and deprivation. Out of these clusters, four clusters were selected for inclusion in a pilot study to assess the feasibility of recruitment, retention and outcome measurement. Feasibility parameters were assessed against stop-go criteria to inform progression to the full cluster randomised trial. Future trial outcome measures were collected at baseline and 3-month follow-up including review of medical records to assess the acceptability and ability of obtaining outcome data in a future evaluation trial.

**Results:** We randomised 32 clusters from 9 catchment areas in 6 NHS Trusts to set up a future multicentre study. We screened 362 people who presented to NHS mental health crisis services in the four identified clusters. A total of 174 service users were identified as potentially eligible for the study with 63 assessed as eligible by the research team. The feasibility study aimed to recruit at least 66% of those deemed eligible, at least 80% of participants retained for interview at 3-month follow-up, and at least 85% retained for the suggested primary outcome obtained from medical records at 3 months. All these criteria were met.

**Conclusions:** A full cluster-randomised controlled effectiveness trial of Open Dialogue within the NHS is feasible in terms of cluster randomisation, participant recruitment and retention, outcome collection, and appropriate delivery of care.

**Key messages regarding feasibility:**

- Whether a cluster randomised controlled trial (cRCT) of OD might be feasibly undertaken within NHS mental health services.
- Feasibility criteria were met with high levels of recruitment and retention
- Large numbers of participants need to be pre-screened in order to reach recruitment targets

## Background

Community mental health services in England, which are provided predominantly by the state funded English National Health Service (NHS), have been the focus of considerable policy development in the past 25 years with the introduction in 2002 of the ‘Functional Team Model’ which promoted the development of a number community teams with specific functions, principally Crisis and Home Treatment Teams, Assertive Outreach Teams and a revised Community Mental Health Team [1]. However, despite initial promise, the care provided by these distinct functional teams has been criticised as being fragmented, with reports of declining continuity of care [2] and dissatisfaction with services [3] [4] [5]. As an approach, Open Dialogue (OD) aims to address the above concerns by providing continuity of care from a single team, with access to a range of treatments including pharmacological, psychological and social interventions [6]. It also places a strong emphasis on working with a service users social network, involving them from the very beginning of an episode of care. OD also has a distinct approach to team working, based on a relational care model, with a clear programme of staff support and supervision [7][8]. OD can be seen as both a treatment intervention and way of organising services, where continuity care is provided by individuals in the multi-disciplinary team from the point of initial crisis through to discharge [9].

A further revision of community mental health policy was produced in 2019 [10] which placed greater emphasis on continuity of care and the involvement of the wider community in the provision of care, both key principles underpinning OD. Whilst non-randomised studies of OD have reported promising outcomes [11] [12], there is limited empirical evidence on how OD itself could be delivered in the UK, and how effective it might be [13][14]. In order to address this limitation in the evidence, the National Institute for Health Research (NIHR), funded a Programme Grant for Applied Research Project, to implement and evaluate the clinical and cost effectiveness of OD in English mental health care services (Open Dialogue: Development and Evaluation of a Social Network Intervention for Severe Mental Illness: ODDESSI, RP-PG-0615-20021). The feasibility study described here is one of the work packages of this programme, which aimed to explore whether a cluster randomised controlled trial (cRCT) of OD might be feasibly undertaken within NHS mental health services.

Cluster randomisation was preferred to enable trial teams to deliver care to a whole locality (and not to only certain individuals within that locality who are allocated to the intervention), in line with ‘usual care’ where people are offered care by a secondary mental health team based on that team’s catchment area. Trial clusters were based on groups of General Practitioner (GP) practices: people in crisis are referred to secondary mental health services through a variety of routes: via self-referral to a mental health ‘triage’ service, via police or ambulance, or via A&E and liaison psychiatry, however the majority of referrals are from primary care, and the majority of people seen in secondary services in the UK are already registered with a primary care GP surgery. The feasibility of this method of participant recruitment, as well as of the proposed application of eligibility criteria and collection of participant interview and medical records-based outcome measures was piloted in two NHS Trusts, informed by the extent to which OD Teams were fully established with OD trained in situ. Finally, the fidelity of service delivery for all OD teams needed to be established.

The specific aims of this feasibility study were:

1. To form and randomise all clusters of GP practices in preparation for a full trial evaluation of the OD approach.
2. To assess the feasibility of service user recruitment and retention in a selected sample of clusters.
3. To assess the feasibility of trial outcome collection in a selected sample of clusters.
4. To establish whether designated OD teams were able to deliver their model of care in preparation for the full trial evaluation.

## Methods

### Restricted randomisation of GP practice clusters

The first aim of the feasibility phase was to randomise all clusters for a future evaluation trial. Clusters here consist of coterminous GP practices from participating mental health trusts. It was important to ensure that the cluster randomisation procedure balanced cluster-level variables believed to be associated with outcomes. These were the catchment area of the Community Mental Health team, ‘average GP list size’ [15] and ‘average IMD deprivation rate 2015’ [16] of the GP practices within each cluster.

The randomisation procedure was as follows: The trial team created clusters of coterminous GP practices. Once an NHS trust and respective catchment areas were recruited the trial team submitted catchment area lists of such clusters to Kings Clinical Trials Unit (KCTU) affiliated statisticians who sequentially randomly allocated clusters to OD or Usual Care at a 1:1 ratio within catchment area strata. Restricted randomisation [17] was used to balance the two continuous aggregated cluster-level covariates GP list size and deprivation rate across the two trial arms, and within and across the catchment area strata. The Chief Investigator and the senior statistician will remain blind throughout any future evaluation study. The junior statistician became partially blind (aware of arms A and B) to facilitate the randomisation. An independent statistician (also facilitating the randomisation procedure), the trial manager, and researchers were unblinded.

### Feasibility pilot sample

Two mental health Trusts were selected to be part of the feasibility pilot: one predominantly urban (NELFT), and one mixed urban-rural (KMPT). Within each of those, two GP practice clusters (from Havering and Canterbury respectively) were selected - one previously allocated to OD and one to Usual Care. Thus, the pilot sample used for feasibility assessment was a selected sample rather than a random sample. Note that GP practice clusters recruited to the feasibility study (but not participants) will also take part in the main trial.

The sample size target for the pilot study was 60 participants from four clusters (15 from each cluster) over a period of 7 months. This was based on wanting to demonstrate the ability to recruit 10% of the sample size required for the main trial over a realistic period.

The pilot study for feasibility assessment received ethical approval from London-Bromley Committee (18/LO/0868) and was conducted in accordance with the Declaration of Helsinki. The CONSORT extension for feasibility studies as well as the CONSORT extension for cluster trials were both considered in the reporting of this manuscript [18] [19].

### Recruitment

#### Mental Health Trusts (future evaluation trial and feasibility study)

In the development of the initial grant application a number of Mental Health Trusts had indicated a willingness to participate in the research programme. Two had already established OD Teams with sufficient staff trained to deliver OD in a locality area, or were ready to establish these teams, and had indicated a willingness to participate in the feasibility study.

#### GP practices (future evaluation trial and feasibility study)

To be considered for inclusion in a trial cluster, GP practices had to:

- Refer a minimum of 10 service users to crisis services per year.
- Have at least 2000 patients registered with the practice.
- Be responsible to serve all residents within their catchment area (e.g. not be primarily a student health service).

Clusters were then formed of 2-3 GP practices which met the criteria above and:

- Were geographically coterminous: have at least one shared catchment area boundary (assessed by looking up the GP practices on a map)
- Referred a total of 65-75 patients per year to crisis services (assessed by adding the number of referrals per practice together-all practices had minimum 10 referrals as per prior practice eligibility).
- Had a clear referral pathway to the community health teams which were the primary provider of crisis and community mental health services for the practices

#### Participants (feasibility study only)

Once the four clusters to be investigated in the feasibility study were formed and pathways into the care provided in the clusters were established, potential participants referred to the secondary mental health teams covered by trial clusters were screened by research staff against pre-determined eligibility criteria; that is they met a) pre-determined initial criteria for a crisis and b) were assessed as being in a mental health crisis at the point of referral. Initial contact with mental health services could be via the established routes for crisis referrals in the services, these included referral to the Emergency Department of the local general hospital and referral to the single point of access for crisis referrals in the participating Mental Health Trust. To participate in the study, participants needed to be assessed as (1) in a mental health crisis (2) be 18 years or above, and (3) able to provide informed consent, or have consent provided by a personal or nominated consultee.

Exclusion criteria were: (1) having a diagnosis of dementia or a learning disability, (2) having a primary diagnosis of substance misuse, (3) having an acquired cognitive impairment, (4) being unable to comprehend both written and verbal English, (5) being under the care of forensic services, (6) being considered too high risk through participation in the study; that is participation in the study is judged by the service users’ clinician to pose potential harm to themselves or the research/clinical team, (7) residing outside their GP catchment area, (8) having no fixed abode, and (9) currently participating in another research trial, which could affect their participation in this trial.

Eligible participants were first asked by a member of their care team whether a researcher could contact them about the feasibility study. Those who agreed were provided with more information and given time (usually 48 hours) to consider their participation. They were then contacted again and asked to provide informed consent and sign a consent form in-person. For those who lacked capacity, consent was sought from either a personal (relative or friend) or nominated consultee (a local senior clinical professional consultee with no other involvement with the trial) according to the Mental Capacity Act [20] [21], with their involvement reviewed again at a time when they regained capacity. Participants received either OD or Usual Care depending on which GP practice cluster they were recruited from, and they continued to receive this care for as long as was required. We also recruited friends or family members of included participants in order to test feasibility of carer recruitment.

### Procedures

#### Assessing the fidelity of delivery of Open Dialogue and Usual Care

OD care takes place primarily through ‘network meetings’, which involve the service user, important others (such as family members or friends), and typically two OD practitioners. Ideally, these practitioners will remain with the network throughout, to provide continuity. The start of each network meeting involves the agenda being set in collaboration with the service user and network members, and with staff working in a collaborative manner to ensure that the key principles of OD are adhered to [9], deploying specific skills to ensure all perspectives are heard and that service users and their network gain a sense of control over their care from the beginning of their care and treatment.

The delivery of OD and of the Community Mental Services (Usual Care) was assessed using a specifically developed fidelity measure, COM-FIDES [21]. This measure was designed to assess the presence of the key components of both OD and Usual Care. This was a requirement of the funder, who wanted to ensure that the care in both arms for the study was delivered to a good standard, thereby ensuring a fair comparison in a future trial. The data for the assessment of fidelity was gained for interviews with a range of staff providing care in both services and by a review of relevant policy and practice documents.

A measure of adherence to OD practice was developed to specifically evaluate the delivery of OD focused on the interactions in OD meetings [23] and assesses the degree to which OD practitioners are delivering theory-specified techniques or methods of intervention [24], based on original work by Olson, Seikkula & Ziedonis [25]. This measure required the audio-recording of network meeting and their rating by a trained panel with significant experience in OD. This scale and the COM-FIDES were used to assesses the extent to which core components of both OD and Usual Care are delivered as intended in teams.

#### Outcome Measures

The baseline (pre-treatment) time point for participants was the date of their crisis referral. The first interview was conducted as close to this date as possible after consent was provided, with the follow-up interview conducted three months later. Interviews occurred either in person (most commonly at the NHS Trust’s research team base or within a participant’s home) or over the telephone. All participants received a £15 shopping voucher after each interview to compensate them for their time.

Medical records were checked to assess the availability of data on service use (including hospitalisation and crisis re-referral), the provision of psychological and pharmacological treatments, and psychiatric status from the crisis point to follow-up at 3 months. The latter would be used to construct relapse and recovery measures for the evaluation trial and so collection and extraction of this information was tested in this study.

Measures of variables that would be further secondary outcomes of the evaluation trial were collected during research interviews or via information extracted from electronic medical records. Outcomes were collected in order to assess acceptability and availability of measures to be used in an evaluation trial. To decrease the burden on participants, half of the outcome measures were used in the first interview and the other half during the follow-up interview at 3 months; see Table 1 for outcome measures collected at each timepoint.

**Table 1.** Outcome measures collected in the feasibility study at each time-point

| **Data source** | **Data collected** | **Time 1**<br>**(baseline)** | **Time 2**<br>**(3 months)** |
| --- | --- | --- | --- |
| **Participant completed measures** | Brief Psychiatric Rating Scale (BPRS) [26] | X |  |
|  | Social Provisions Scale (SPS) [27] | X |  |
|  | Lubben Social Network Scale (LSNS-6) [28] | X |  |
|  | Health related quality of life (EQ-5D-5L) [29] | X |  |
|  | Client Satisfaction Questionnaire (CSQ) [30] |  | X |
|  | Questionnaire about the Process of Recovery (QPR) [31] |  | X |
|  | Client Services Receipt Inventory (CSRI) [32] |  | X |
|  | Modified Dyadic OPTION scale [33] |  | X |
| **Family /carers** | Burden Assessment Scale (BAS) [34] | X |  |
|  | Client Satisfaction Questionnaire (CSQ) [30] |  | X |
| **Medical note review** | Relapse and recovery summary [35] |  | X |
|  | Admission | X |  |

#### Feasibility parameters

Six ‘progression criteria’ were agreed with the programme funders (NIHR). The first four relate to the feasibility study conducted in two catchment areas/four clusters and are measured at a participant level, and the remaining criteria relate to all proposed NHS Trust trial sites and are assessed at a site level.

1. Recruitment of 10% of the proposed evaluation trial participant sample over a 7-month period (participant-level)
2. Consent rate of at least 66% (participant level)
3. Retaining 80% of participants at 3-month follow-up (participant level)
4. Proposed primary outcome data (relapse) collection from 85% of recruited participants from medical notes (participant level)
5. OD teams operating within agreed protocols (site level)
6. Usual Care and OD teams’ fidelity and OD adherence (site level)

Relevant feasibility parameters (e.g. number recruited, percentage retained) were defined and progression criteria are described as ‘met’ if the actual number falls within 15% of the predefined criteria.

To satisfy parameters 5 and 6, teams were required to achieve adequate fidelity and adherence scores prior to commencement of participant recruitment (both prior to the feasibility trial for the two participating teams, and prior to the main trial for all teams).

#### Analysis

As recruitment and randomisation of all clusters for the future evaluation trial was part of the feasibility phase the full procedure will be described in the Results Section.

Feasibility parameters are mainly assessed using figures from the CONSORT diagram. During completion of the study, it became clear that the number of participants who consented should be presented in two different ways. The number of participants who consented out of those who were deemed to be eligible in pre-screening and also the number who consented out of those deemed eligible after the full screening. We calculated proportion consented of those eligible (at both stages) overall and by cluster; recruitment rate (number of participants recruited) overall and by cluster and follow-up rates at 3 months (participants who have completed measures out of those recruited). Feasibility parameter estimates were accompanied by 95% confidence intervals to provide a measure of precision; for proportions these were generated using the binomial exact distribution with the cii proportions command in Stata. The cii means command with the Poisson distribution option was used for the calculation of confidence intervals for rates.

## Results

### Cluster formation and randomisation

UCL research staff liaised with senior clinical and managerial staff and staff in the participating Trusts’ data management services to provide a list of GP practices, and the number of crisis referrals from each GP practice to the mental health service in the previous 1- or 2-year period.

In order to form clusters for a catchment area, an initial consideration was the capacity of the OD Team to provide sufficient input to support the feasibility study, further considerations were the likelihood of generating sufficient referrals to support the study and to form a coterminous cluster of 2-4 practices.

Once one cluster was formed, the process was repeated to attempt to form the required number of clusters meeting criteria using the remaining GP practices. This process was iterative: if sufficient clusters could not be formed then GP practices could be moved to a different cluster until all criteria were met and sufficient clusters formed.

Once a list of proposed clusters was established for each catchment area, this was reviewed in collaboration with senior clinical and managerial staff in the NHS Trusts to confirm their willingness and ability to deliver OD to any of the clusters established. Once confirmed by senior clinical and managerial staff the clusters were considered finalised and ready for randomisation. Information was then collated to describe the clusters: number of full-time equivalent GPs, number of GP partners, average GP list size and average IMD deprivation rate 2015 to be used within the randomisation procedure.

**Figure 1.**
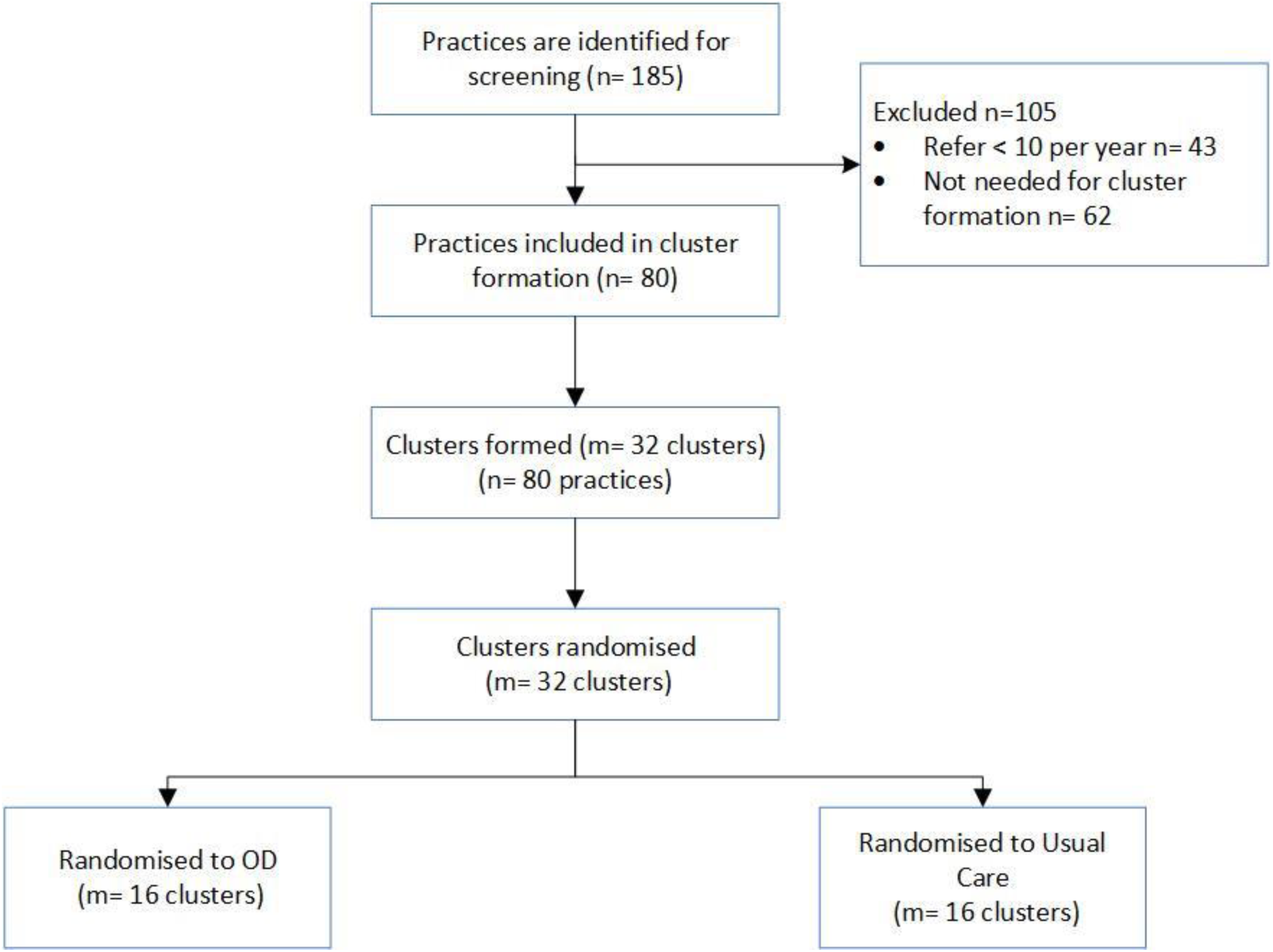
shows the flow of practices and the process that was undertaken to form the clusters to be randomised. Of the 185 practices that were identified, 80 were eligible and went on to form our 32 trial clusters. All 32 were randomised, 16 to each arm.

### Feasibility study

Cluster characteristics can be seen in Table 2. We will be reporting data split by catchment area, not by arm, to maintain blinding for the remainder of the ODDESSI programme. Each catchment area here contains one intervention cluster and one Usual Care cluster.

**Table 2.** GP practice cluster characteristics

|  | **Havering M=2** | **Canterbury M=2** | **Overall M=4** |
| --- | --- | --- | --- |
|  | **Mean** | **Mean** | **Mean (SD)** |
| **Number of equivalent full time GPs** | 14.5 | 11.0 | 12.8 (2.9) |
| **Number of registered GP partners** | 11.0 | 7.5 | 8.7 (2.5) |
| **Rural or Urban classification *N*** |  |  |  |
| *Rural* | 0 | 1 | 1 |
| *Urban* | 2 | 1 | 3 |
| **Average GP list size** | 8195.0 | 12832.0 | 10513.5<br>(3642.7) |
| **Average IMD score** | 15.6 | 17.5 | 16.6 (2.7) |

Recruitment of participants from the four chosen clusters from these areas took place between 30/07/2018 and 18/02/2019, with 59 participants being recruited. Figure 2 shows participant flow through the trial. The most common reason for participants being ineligible was that they were not in crisis, and of those seemingly eligible from the initial pre-screening the most common reason for not going on to consent was that they could not be contacted. We were only able to recruit a carer/family member for 24 participants.

**Fig 2.**
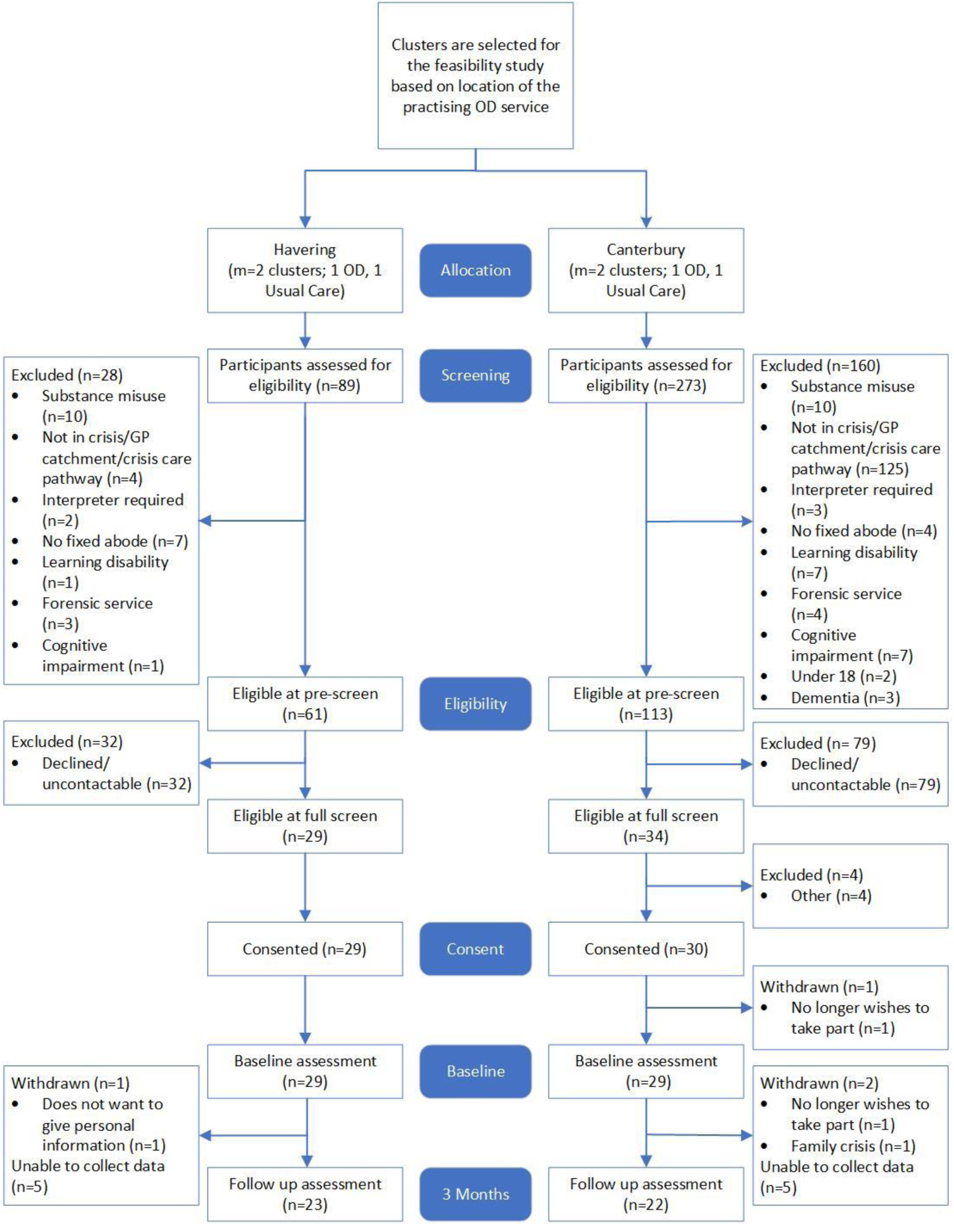
The flow of participants through the feasibility study

The majority of the participants were female (57.6%), heterosexual (76.3%), and born in the UK (89.8%). All baseline characteristics are shown in Table 3 and were similar between catchment areas.

**Table 3.** Summaries of socio-demographic variables for study participants

| Baseline demographics |  | Havering<br>N=29<br>N (%) | Canterbury<br>N=30<br>N (%) | Overall<br>N=59<br>N (%) |
| --- | --- | --- | --- | --- |
| **Age at consent** | *mean (SD)* | 37.0 (14.4) | 38.6 (12.6) | 37.8 (13.5) |
| **Gender** | *Female* | 15 (51.7) | 19 (63.3) | 34 (57.6) |
|  | *Male* | 14 (48.3) | 9 (30.0) | 23 (39.0) |
|  | *A gender identity not listed* | 0 (0.0) | 1 (3.3) | 1 (1.7) |
|  | *Missing* | 0 (0.0) | 1 (3.3) | 1 (1.7) |
| **Sexual orientation** | *Heterosexual/Straight* | 24 (82.8) | 21 (70.0) | 45 (76.3) |
|  | *Other* | 5 (17.2) | 7 (23.3) | 12 (20.3) |
|  | *Missing* | 0 (0.0) | 2 (6.6) | 2 (3.4) |
| **Country of birth** | *UK* | 26 (89.7) | 27 (90) | 53 (89.8) |
|  | *Other* | 3 (10.3) | 2 (6.6) | 5 (8.5) |
|  | *Missing* | 0 (0.0) | 1 (3.3) | 1 (1.7) |
| **Education** | *Primary education or less* | 2 (6.9) | 0 (0.0) | 2 (3.4) |
|  | *Secondary education* | 18 (62.1) | 15 (50.0) | 33 (55.9) |
|  | *Tertiary/ further education* | 7 (24.1) | 11 (36.7) | 18 (30.5) |
|  | *Other* | 2 (6.9) | 3 (10.0) | 5 (8.5) |
|  | *Missing* | 0 (0.0) | 1 (3.3) | 1 (1.7) |
| **Employment** | *Employed* | 11 (37.9) | 8 (26.7) | 19 (32.2) |
|  | *Unemployed* | 7 (24.1) | 7 (23.3) | 14 (23.7) |
|  | *Full time education* | 1 (3.4) | 0 (0.0) | 1 (1.7) |
|  | *Not working due to disability* | 9 (31.0) | 12 (40.0) | 21 (35.6) |
|  | *Looking after child or home* | 1 (3.4) | 0 (0.0) | 1 (1.7) |
|  | *Retired* | 0 (0.0) | 2 (6.7) | 2 (3.4) |
|  | *Missing* | 0 (0.0) | 1 (3.3) | 1 (1.7) |
| **Marital status** | *Single* | 17 (58.6) | 15 (50.0) | 32 (54.2) |
|  | *Married/cohabiting/civil partnership* | 8 (27.6) | 12 (40.0) | 20 (33.9) |
|  | *Separated/divorced* | 4 (13.8) | 2 (6.7) | 6 (10.2) |
|  | *Missing* | 0 (0.0) | 1 (3.3) | 1 (1.7) |
| **Housing status** | *Owner* | 6 (20.7) | 5 (16.7) | 11 (18.6) |
|  | *Renter* | 8 (27.6) | 6 (20.0) | 14 (23.7) |
|  | *Live with relatives* | 10 (34.5) | 10 (33.3) | 20 (33.9) |
|  | *Social/council housing* | 4 (13.8) | 7 (23.3) | 11 (18.6) |
|  | *Care home* | 1 (3.4) | 0 (0.0) | 1 (1.7) |

Feasibility parameter estimates are shown in Table 4. In terms of our four participant level feasibility outcomes, the overall consent rate of those deemed eligible through pre-screening was 34% and these consents were received at a rate of 8.4 per month over the 7 months recruitment period. The consent rate for those eligible after full screening was 93.7% at a rate of13.4 per month. There was a high retention percentage with only 4 out of 59 participants withdrawing from the study, 3 due to participants no longer wishing to take part in the study and 1 due to participant distress. Collection of data from medical notes review on admission and re-referral information was very high with data being extracted and collected from 96.6% of consented participants. Availability of questionnaire outcome was high at baseline (>95%) but there was some attrition observed at 3 months follow-up (up to 26% for the worst recorded OPTION measure). There were no adverse events recorded throughout the study.

**Table 4.** Feasibility results

| **Feasibility parameters** | ***Proportion/rate [CI]*** | ***Numbers used*** |
| --- | --- | --- |
| **Proportion consented of those eligible from pre-screening (%)** | 34.0 [26.9, 41.5] | 59/174 |
| **Proportion consented of those eligible from full screening (%)** | 93.7 [84.5, 98.2] | 59/63 |
| **Recruitment rate overall (*rate*)** | 8.4 [6.4, 10.9] | 59 in 7 months |
| **Recruitment rate by cluster (*rate*)** |  |  |
| *Cluster 1* | 3.7 [1.8, 6.6] | 11 in 3 months |
| *Cluster 2* | 6.0 [3.6, 9.5] | 18 in 3 months |
| *Cluster 3* | 2.2 [1.2, 3.7] | 13 in 6 months |
| *Cluster 4* | 3.1 [1.8, 4.9] | 17 in 5.5 months |
| **Proportion of baseline outcome completion (%)** |  |  |
| *SPS* | 98.3 [90.9, 100.0] | 58/59 |
| *LSNS6* | 98.3 [90.9, 100.0] | 58/59 |
| *BPRS* | 96.6 [88.3, 99.6] | 57/59 |
| *EQ5D* | 98.3 [90.9, 100.0] | 58/59 |
| **Proportion of follow up outcome completion (%)** |  |  |
| *Admission* | 96.6 [88.3, 99.6] | 57/59 |
| *Rereferral* | 96.6 [88.3, 99.6] | 57/59 |
| *CSQ-8* | 76.3 [63.4, 86.4] | 45/59 |
| *CSRI* | 76.3 [63.4, 86.4] | 45/59 |
| *OPTION* | 74.6 [61.6, 85.0] | 44/59 |
| *QPR* | 76.3 [63.4, 86.4] | 45/59 |

Prior to participation in the feasibility recruitment, the OD and Usual Care teams for those four clusters met criteria for fidelity on the COM-FIDES measure [21], and the OD teams provided audio-recordings of network meetings which met adherence criteria [22], and operational protocols by which their services were being delivered. All further teams met these criteria prior to recruitment of participants to the main trial.

## Discussion

This study sought to test the feasibility of conducting a cRCT assessing the effectiveness of OD within the NHS. The results indicate that it was feasible to recruit and retain participants in four pilot clusters, and that the proposed outcome measures were largely acceptable as seen by the feasibility criteria being met.

There were several strengths of this study. We utilised a pragmatic design which was based in NHS mental health services. Therefore, a key part of this feasibility study included successfully establishing OD teams in the Trusts who would take part in the main trial. This involved significant collaboration with Finnish and international OD practitioners and trainers, all of which was successfully completed in the required timeframe.

Despite this study confirming the feasibility of a full evaluation trial, some minor adjustments will need to be made. The information gained on the consent numbers shows the importance of pre-screening a large number of participants prior to full formal eligibility screening in order to recruit the number of participants needed due to large numbers of seemingly eligible participants being uncontactable. Feedback from participants about the difficulty of being able to make a face-to-face appointment for consent and follow-up assessments also means that there are plans to make these contacts by telephone where appropriate. Similarly, recruiting family members/carers took researchers significant time and so this process needs to be streamlined for the main trial.

It was not possible for the researchers conducting participant interviews to remain blind to intervention allocation. Their roles required liaising with service users clinical teams, accessing their medical records, speaking with them about their care, and visiting them at home, all of which are unblinding. Keeping these embedded researchers blind during the evaluation trial appears to be impractical and only statisticians and the Chief Investigator, and the panel assessing the primary outcome, will remain blind for the main trial. Furthermore, establishing a diagnosis using the ICD criteria [36] at baseline was found to be difficult because it frequently delayed the time to recruit participants. Instead, the A-C rating on the UK Mental Health Triage Scale [37] was deemed appropriate for classifying mental health crisis on review of the investigators and research team and will be used to assess the possible presence of a mental health crisis in the evaluation trial.

After completing this study, it was decided that previous receipt of the OD intervention should also be added as an extra exclusion criterion for participants of the evaluation trial. Approaching the start of the ODDESSI programme, some of the KMPT and NELFT teams already trained in OD had been delivering the intervention for a few years, and so including this as an exclusion means that anyone who had previously used the service could not now enter into the trial and potentially bias the results.

A limitation of this feasibility study is that it only followed up participants for three months, and so it is difficult at this point to infer the full attrition rates of the evaluation trial, which will include a two-year follow-up. The success of extracting data from participant electronic health records for the potential primary outcome of the main trial is somewhat protective against the potential impact of participants being lost to follow-up.

## Conclusion

This feasibility study has informed the design of the largest multicentre cRCT comparing OD to Usual Care within the NHS to date. With a few minor adjustments, the results indicate that OD teams can be established, and participants can be successfully recruited, consented and retained, and that the outcome measures are acceptable.

## Declarations

### Ethics approval and consent to participate

This study for feasibility assessment received ethical approval from London-Bromley Committee (18/LO/0868)

### Consent for publication

Not applicable

## Data Availability

The datasets generated and/or analysed during the current study are not publicly available due to the programme still being ongoing but will be available from the corresponding author on reasonable request once completed.

## Authors’ contributions

The study was conceived by SP who was the Chief Investigator. The first draft of the manuscript was written by SP, KJ, KC, GP and SL. The data was analysed and interpreted by KJ and SL. SH was responsible for recruitment, data acquisition and management and study administration. YI was the site manager for KMPT and involved in recruitment. CK provided strategic oversight to implementation in KMPT and contributed to study design and development of intervention. JO was the clinical lead and Principal Investigator for KMPT. RR was the Principal Investigator for NELFT, Open Dialogue trainer and national clinical lead for the study. All authors read and approved the final manuscript.

## Acknowledgements

We would like to thank the clinical teams and the patients from our participating trusts for contributing their time to the recruitment and follow-ups. We are grateful for the contributions of KMPT Researcher Marcus Colman, whose work during the set-up and conduct of this work was key to its successful delivery. This work was part of a larger NIHR programme grant for which several further co-applicants contributed in terms of planning and design, including: Jerry Tew, Tim Weaver, Sonia Johnson, Stephen Morris, Sarah Carr, Corrine Hendy, Mark Hopfenbeck, Douglas Ziedonis, and John Brouder.

